# An estimation of the health-cost of unfilled medical positions in Malawi: A Thanzi La Onse Mathematical Modelling study

**DOI:** 10.64898/2026.05.25.26353761

**Authors:** Aksayaa Perinpakumar, Bingling She, Tara Mangal, Sakshi Mohan, Martin Chalkley, Tim Colbourn, Joseph H. Collins, Matthew M. Graham, Eva Janoušková, Dominic Nkhoma, Pakwanja D. Twea, Andrew N. Phillips, Paul Revill, Asif U. Tamuri, Joseph Mfutso-Bengo, Timothy B. Hallett, Margherita Molaro

## Abstract

**Background:** Malawi’s healthcare system faces strain due to an insufficient number of healthcare workers (HCWs). The number of HCWs currently employed falls below the Malawian government’s own facility-based staffing standards, which are known as the *establishment target*. While vacancy rates from this target have been estimated, the health consequences of this workforce gap on the population have not.

**Methods:** This study quantifies the health-cost of unfilled establishment HCW positions using the Thanzi La Onse (TLO) model, an “all diseases – whole healthcare system” individual-based model, which self-consistently accounts for the dynamics between health system constraints and population health. We constructed two staffing scenarios: one (*Current)* in which the currently employed staff are represented, and another (*Target)* where all positions planned under the establishment target are filled. Using the TLO model, we then estimate the health impact of filling all establishment positions as the difference in the Disability-Adjusted Life Years (DALYs) incurred between the two scenarios.

**Results:** Our results indicate that fulfilling Target positions could reduce the health losses by 13.6% (43.1 million DALYs averted, 95% CI: 40.8–48.6) over the projection period. The largest proportional reductions are for DALYs caused by HIV/AIDS (41%), tuberculosis (26%), and malaria (24%) compared to the Current provision.

**Conclusions:** The analysis shows the potential health benefits associated with increasing the fulfilment of establishment positions in Malawi and offers key quantifications for policymakers as they strive to achieve Universal Health Coverage.

**Author Summary:** Malawi’s current healthcare workforce is significantly smaller in number than the *establishment* targets set out by the Ministry of Health. Our research uses an individual-based epidemiological and healthcare system model, *Thanzi La Onse*, to examine the consequences of filling all the target healthcare worker positions. We find that the healthcare system could reduce the overall disease burden by an extra 43.1 million DALYs (13.6% reduction) between 2023 and 2040, with the greatest reductions in the burden of AIDS, tuberculosis, and malaria. This modelling approach accounts for other barriers to healthcare delivery, such as healthcare-seeking habits, consumables and equipment supply, diagnostic accuracy, and treatment efficacy. Our findings, therefore, show that in Malawi’s healthcare system, every unfilled healthcare worker post represents an important lost opportunity for population health gains.

## Introduction

A healthcare system’s overall impact on the health of the population it serves is driven, in part, by the availability, skills, motivation, and appropriate deployment of healthcare workers (HCWs)^1^. Lack of HCWs means loss of population health^2–4^. Hence, the United Nations’ Sustainable and Millennium Development Goals (SDGs and MDGs) have explicitly prioritised increased health financing alongside the recruitment, development, training, and retention of the healthcare workforce^5,6^. WHO has identified 55 countries, including Malawi, that it believes lack sufficient HCWs to achieve the UN Sustainable Development Goal of UHC by 2030^1^.

Malawi has achieved remarkable health progress within the last two decades, including an 18-year increase in life expectancy at birth between 2000 and 2021^7^. However, multiple health challenges remain, such as high prevalence of infectious disease, high maternal mortality and infant mortality, as well as growing non-communicable disease burden^8–16^. The demand for healthcare exceeds the country’s capacity, constrained by limited resources, infrastructure, and personnel^17^. WHO benchmarks recommend at least 4.45 doctors, nurses, and midwives per 1,000 population to achieve universal health coverage. Malawi falls well below this threshold (an estimated 0.56 per 1000, in 2022)^7^. Although the government of Malawi sets out targets for the number of HCWs (of different types) that should be employed in each healthcare facility type, known as the *establishment*, many HCW positions remain unfilled^17–21^. In a constrained and over-burdened healthcare system, these unfilled positions potentially reflect a significant, but as of yet unquantified, health opportunity cost.

Healthcare workforce pipeline models and health workforce planning (supply-demand) models are valuable tools for planning future healthcare workforce scenarios^22^. Several have been applied to Malawi, for example, the Workforce Optimisation Model (WFOM)^23^ which was developed by the Clinton Health Access Initiative (CHAI) and the Zambian Ministry of Health (MoH) in 2009, as well as the Workload Indicators of Staffing Need (WISN)^20,24^ method developed by WHO as an approach for human resources for health (HRH) planning utilised in multiple countries, Malawi among them. These models have contributed to estimating healthcare workforce requirements by incorporating service delivery needs, health worker time-and-motion studies, and staffing norms. They are, however, limited in their capacity to investigate the possible health consequences of different health workforce allocation scenarios. In contrast, the *Thanzi La Onse* (TLO^25^) model integrates epidemiological and demographic dynamics with healthcare system usage and constraints^25^. This allows for both the forecasting of workforce requirements and the estimation of the consequences for population health from different levels of HCW staffing levels, while accounting for shifts in disease patterns, patient behaviour, and healthcare-seeking trends resulting from the extent to which available HCW capabilities can meet demand.

This article aims to use the TLO model to quantify the health loss associated with HCW vacancies in Malawi by evaluating the health losses that could be averted if all vacant establishment positions were to be filled. In particular, we use the TLO model to quantify under different healthcare workforce scenarios, (i) the difference in total and cause-specific Disability-Adjusted Life Years (DALYs) occurring; and (ii) the effect on healthcare services delivered.

## Methods

The TLO model is an individual-based model that captures multi-disease population health, health-related risk factors, healthcare-seeking behaviour, and the interaction of individuals who seek care with the healthcare system. In particular, the ability of the healthcare system to meet the demand for care is constrained by the availability of required resources, including HCWs, consumables, medical equipment, and beds. This setup allows the model to simulate how healthcare system constraints influence individual health outcomes. The detailed and mechanistic representation of the healthcare system–epidemiology interaction is a distinctive feature of the TLO framework. It has been described in detail elsewhere^26^ but is briefly summarised below.

The TLO explicitly models the leading causes of DALYs and deaths in Malawi, including HIV/AIDS, TB, malaria, conditions related to maternal and newborn health, acute lower respiratory infections, measles, childhood diarrhoea and stunting, road traffic injuries, cardio-metabolic disorders, chronic obstructive pulmonary disease, depression, diarrhoea, epilepsy, and cancers^7^. Once a simulated individual has developed symptoms, the question of whether individuals initially seek care or not is assumed to be dependent on the lifestyle and demographic features they possess, alongside the nature of the symptoms they are experiencing (see ^26,27^ and references therein).

Once individuals have sought care in the model, care is provided via a ‘Health System Interaction’ (HSI) event. In total, 49 types of health service interactions (“HSIs”) are captured by the model, with each type of HSI specifying: (i) the level at which care should be delivered, (ii) the minutes of HCW patient-facing time required for delivery; (iii) the proportion of that service time attributed to each cadre (nurses, pharmacists, clinicians, etc); and (iv) consumables, equipment, and beds required. These requirements vary by facility type, reflecting the different provisions of health posts, health centres, community hospitals, district hospitals, and central hospitals. Consequently, HSIs serve as indicators of the demand for healthcare services and the workforce needed to deliver care.

### Representation of the Health Service Delivery in the TLO Model

In the TLO model, health services in Malawi are recognised as being delivered through a four-tier system - community, primary, secondary and tertiary levels (see ^17^ and ^26^ for further explanation). Each facility level in each district is assumed to have a finite number of resources to treat patients (see^17^). In particular, with regard to human resources for health, this includes an overall amount of patient-facing time available per day per HCW cadre in each simulated facility. HSIs are therefore scheduled against available HCW working time: when demand exceeds capacity, some are not delivered or are delayed until patients are assumed to desist (outlined within the overview paper^26^). Thus, unmet healthcare need arising from insufficient HCW time is captured dynamically within the model. Similarly, assumed consumable availability reflects the observed availability^28^ per consumable type and facility, such that if a required consumable is not available at the time of HSI delivery, the success of the HSI and therefore its impact on the patient is compromised.

### Analysis Design

We constructed two scenarios of HCW staffing patterns:

- The *Current* scenario, which reflects the staffing levels observed in the year 2019^29^.
- The *Target* scenario assumes that the Government of Malawi’s establishment targets for staffing levels in each type of healthcare facility are fully met. In addition, this scenario assumes (i) a redistribution of mental health staff from regional hospitals so that every district has at least one mental health worker, with staff divided between primary hospitals (around 48%) and district hospitals (around 52%); and (ii) allocation of community health assistants (DCSAs) to cover underserved areas such as Likoma district. These adjustments make staffing levels more consistent with the expected demand for services across the health system^20,30^; the parameterisation for human resources for health (HRH) in the two scenarios used data from the Detailed Annex for the Health Workforce Interventions in Malawi’s HSSP III (2023-2030)^26^ and the data used for research purposes were accessed on 17/02/2025.

According to the locally defined establishment target for facility staffing, current staffing levels reveal gaps across many professional categories. The differences in staffing levels between the two scenarios are summarised in Fig. 1. The highest percentage of posts unfilled (vacancies) is seen in: Mental Health Care Specialists (88.6%), Dental Specialists (80.3%), Pharmacists (61.0%) and Radiographers (54.4%), where the percentages indicate how much of the Target positions remain unfilled. In contrast, for DCSAs, the Current staffing is already at the Target level. This reflects a closer alignment between current and Target numbers for DCSAs under existing programmes, unlike other cadres that remain heavily under-recruited.

**Figure 1.**
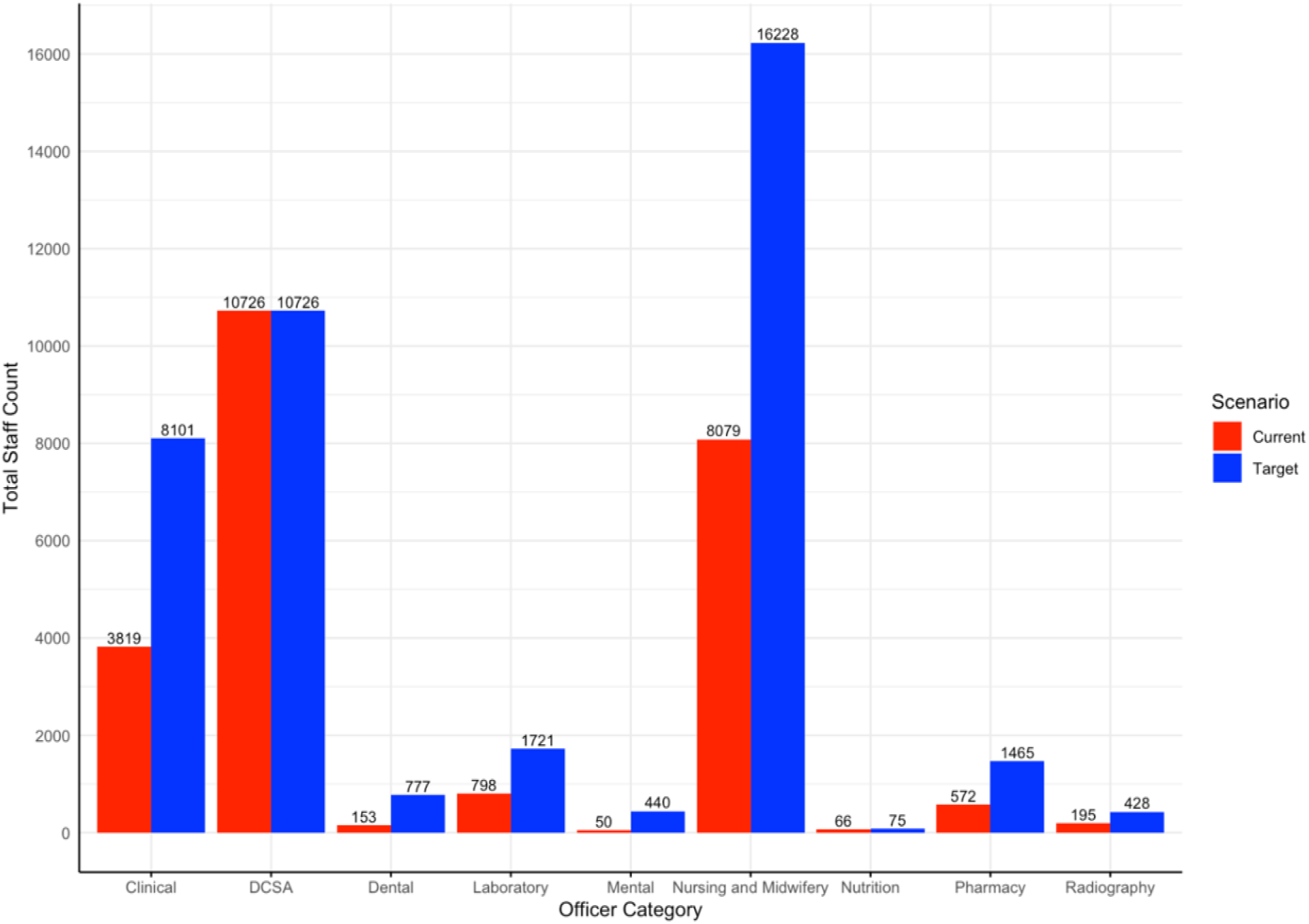
Bar graph showing the distribution of total staff counts across different officer categories within Malawi’s healthcare system.

We simulate both staffing scenarios using the TLO model for the period of 1 January 2023 to 31 December 2040. A time horizon to the end of 2040 allows for insights to be drawn from the trajectory over time of unmet needs beyond the SDGs and Malawi’s HSSP III milestones, and the potential health cost of continued under-staffing, helping policymakers anticipate and plan for challenges beyond short-term strategy cycles. We assume no differences between the two scenarios with regard to all remaining elements of the healthcare system (such as consumable availability, hospital beds, and equipment (e.g., diagnostic machines)). We performed 10 simulations for each scenario, and then computed summary statistics on these results to estimate the difference between the two scenarios, with respect to the total DALYS incurred, the cause of the DALYs incurred, and the number of HSIs that were delivered per cause. The model is run using Python language version 3.8; all source code is available on GitHub^25^.

## Results

The projected median DALYs incurred each year from 2023 to 2040 under both scenarios are presented in Fig. 2. The Current scenario (red line) consistently shows higher DALYs across the entire period compared to the Target scenario (blue line). This shows that with all establishment positions filled (in the Target scenario), the overall health burden would be consistently lower throughout 2023–2040. Across the whole period, the Current scenario accumulates a median of 317.7 million DALYs (95% CI: 315.3–323.7), while the Target scenario accumulates 274.8 million DALYs (95% CI: 273.3–278.0). This represents a difference of 43.1 million DALYs averted (95% CI: 40.8–48.6), equivalent to a 13.6% reduction in health burden incurred in the whole population by 2040.

**Figure 2.**
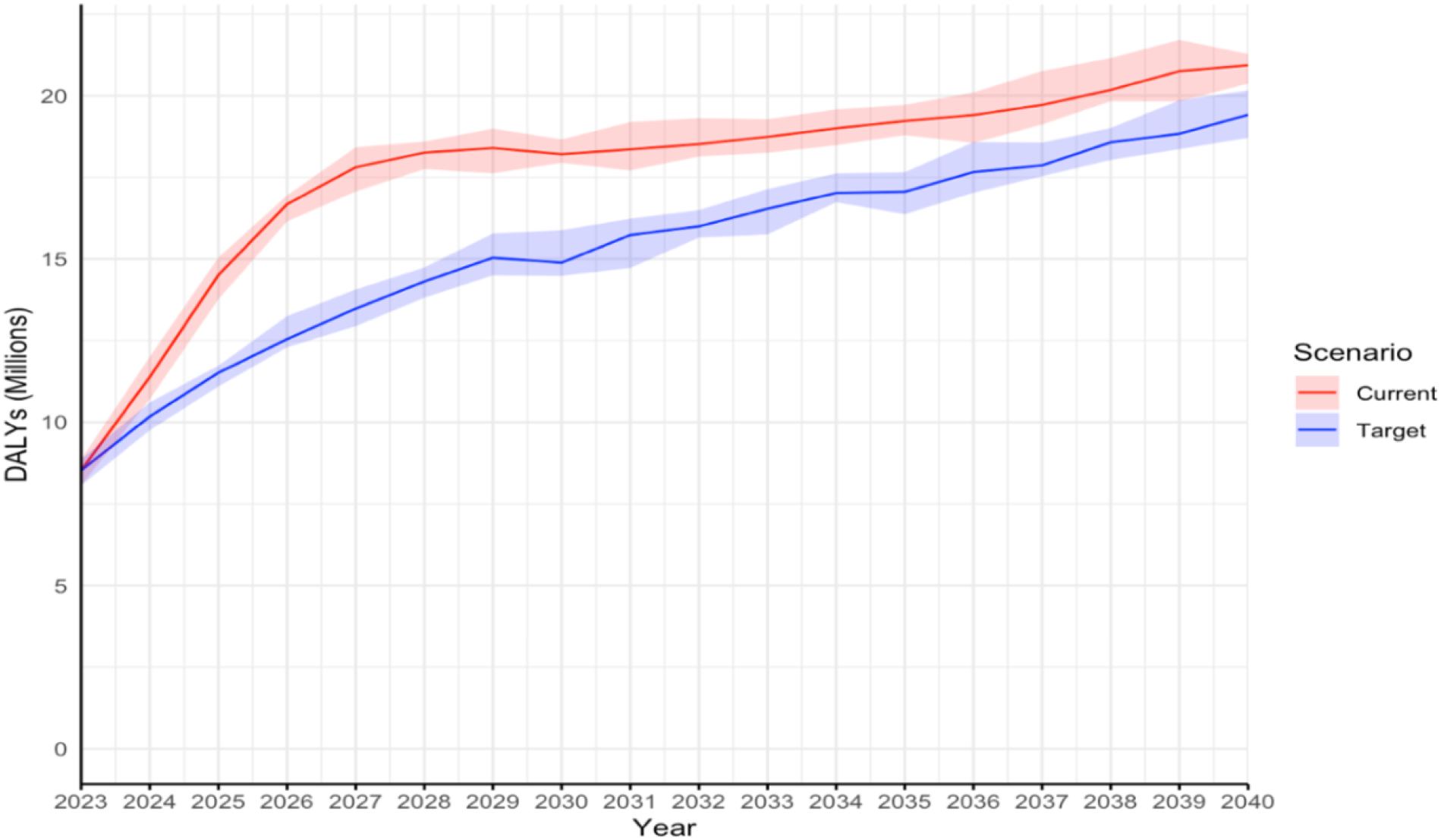
Projected median DALYs accrued each year over time from 2023 to 2040. Shaded areas represent the 95% interval for each statistic.

The DALYs incurred in the period 2023–2040 are broken down by their cause in Fig. 3. For AIDS, in the Target scenario, cumulative DALYs are 41% lower than in the Current scenario (55.3 million in the Current vs. 32.6 million in the Target). TB (non-AIDS) is 26% lower (16.4 million to 12.2 million), Malaria is 24% lower (20.5 million to 15.6 million), and Measles is 31% lower (10.6 million to 7.3 million). Lower respiratory infections also decline by 8% (60.2 million to 55.3 million) between the two scenarios, and Neonatal disorders are lower by 9% (41.6 million to 37.9 million). In contrast, the difference in health losses to other causes does not differ meaningfully between the scenarios: depression/self-harm (3% higher), transport injuries (0.4% lower), and stroke (0% change). Overall, there is no meaningful difference between the scenarios for these causes.

**Figure 3.**
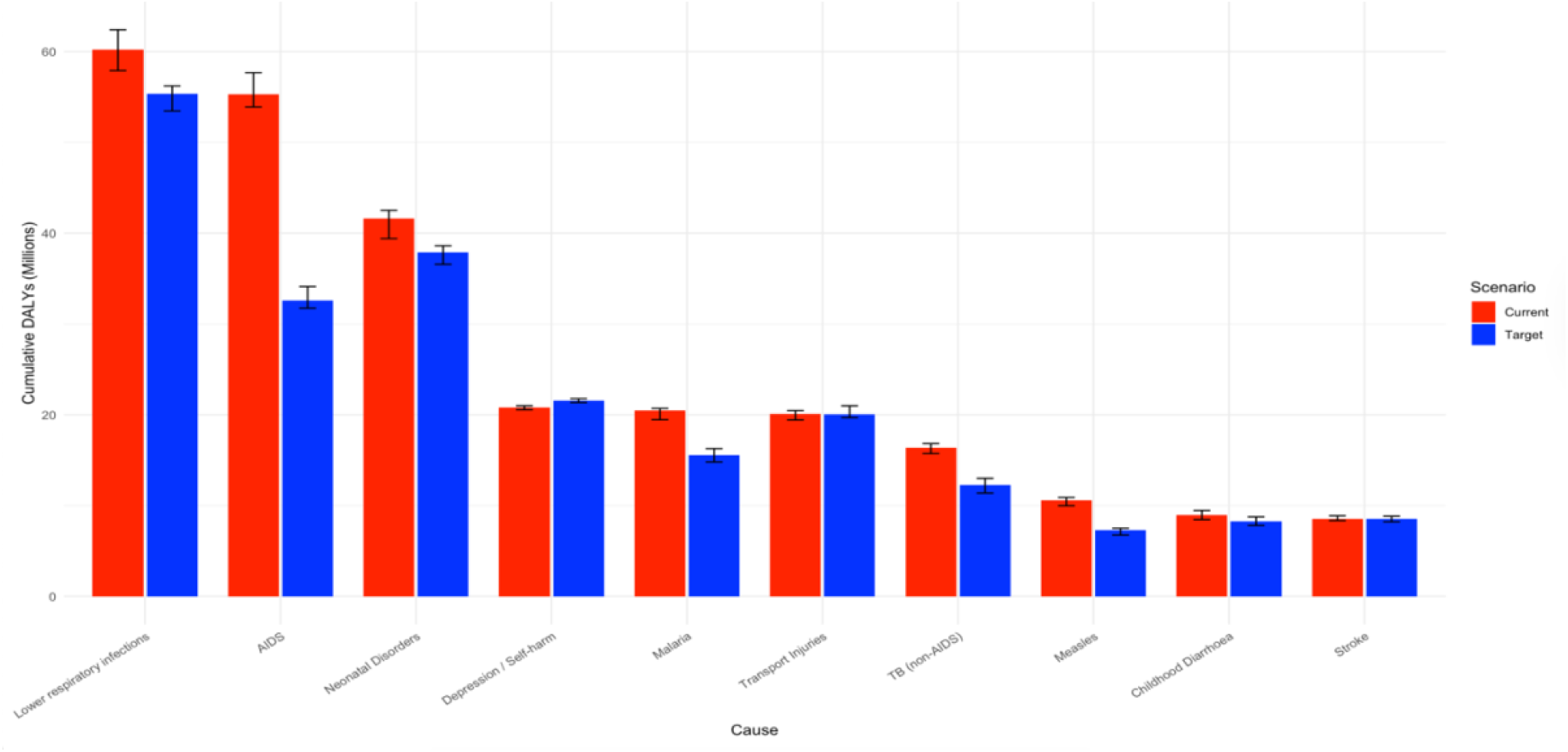
Bar chart displaying the top 10 causes of cumulative DALYs by 2040 for both Current and Target scenarios. Error bars indicate the 95% confidence CI.

Across almost all causes, the Target staffing scenario enables higher delivery of health service interventions (HSIs) than the Current scenario (Fig. 4), given the log scale, even modest horizontal shifts, correspond to large absolute differences in service volume. The largest absolute increases occur in high-volume services. Contraception delivery rises from approximately 454 million appointments (95% CI 444–461 million) under the Current staffing to 600 million (583–607 million) under the Target scenario. Similarly, EPI services increase from 74 million (73–74 million) to 184 million (182–184 million), and TB (non-AIDS) more than doubles, from 53 million (53–54 million) to 138 million (137– 138 million). Moderate gains are observed for several mid-volume causes, including cardiometabolic disorders (61 million vs. 37 million), antenatal care (20 million vs. 3 million), diarrhoea (19 million vs. 9 million), depression/self-harm (13 million vs. 2 million), epilepsy (20 million vs. 13 million), lower respiratory infections (9 million vs. 6 million) and Transport injuries show minimal change. Overall, the Target scenario consistently enables higher cumulative delivery of HSIs across almost all causes, with the most substantial gains in contraception, EPI, TB, HIV, and malaria.

**Figure 4.**
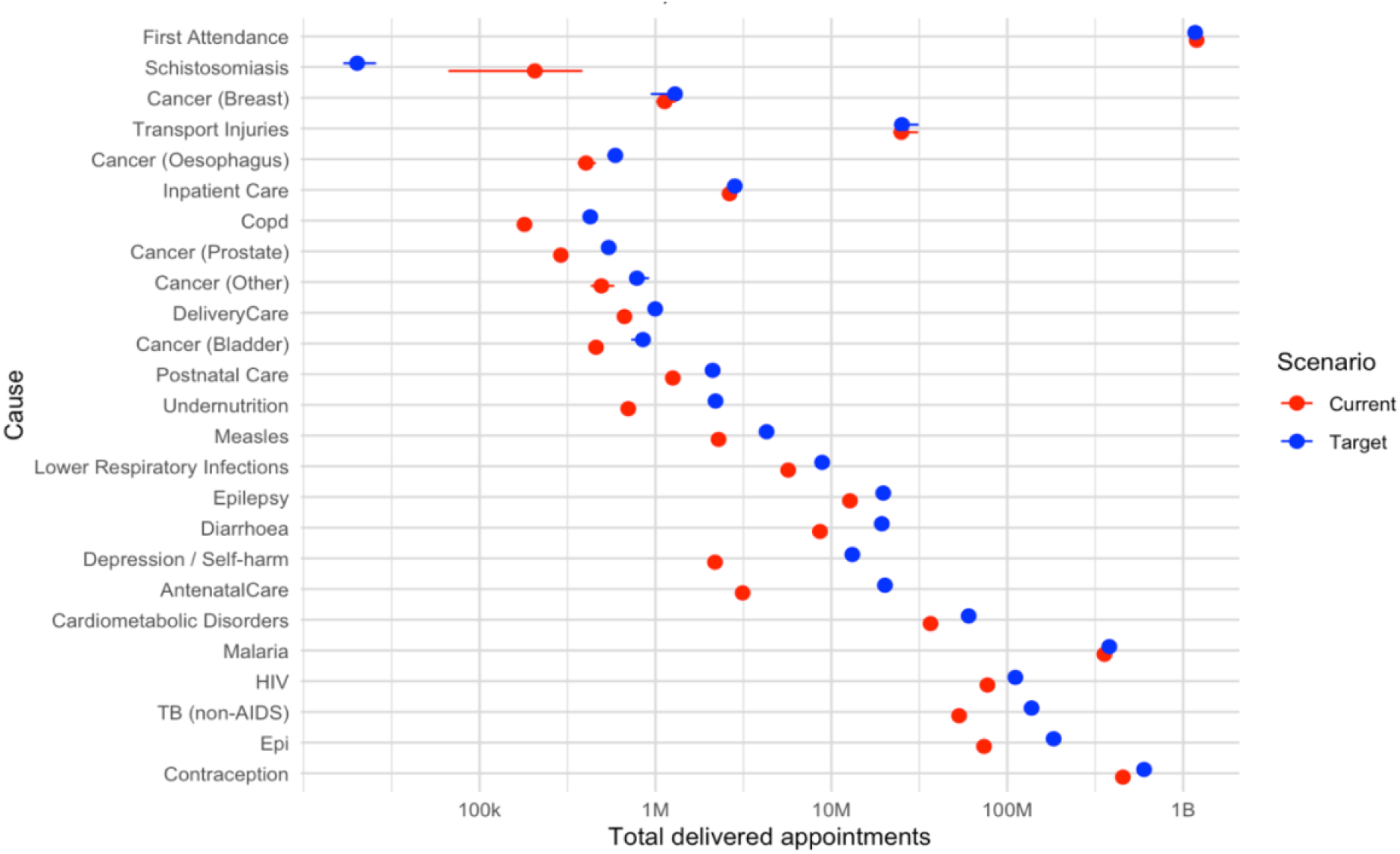
Points show median cumulative delivered HSIs for each cause under each scenario; horizontal bars indicate 95% CIs. The x-axis is plotted on a log_10_ scale (millions, M) to accommodate differences in service volume across causes.

## Discussion

If all healthcare worker positions defined in the Government of Malawi’s establishment were filled, it could lead to a 13.6% overall reduction in DALYs over 2023–2040, equivalent to 43.1 million DALYs averted under the assumption of current resource allocation across diseases. Among the top 10 causes of DALYs, the largest difference would be for AIDS (41% lower with all positions filled), TB (26% lower), Malaria (24% lower), and Measles (31% lower): this underscores that scaling up workforce capacity can alleviate health burdens for high-priority conditions, including infectious diseases that rely heavily on continuous service delivery Expanded workforce availability substantially increases the system’s overall capacity to deliver care over time (Fig. 4), providing a clearer indication of the scale of services that can be supported for planning, budgeting, and forecasting of commodities and workforce requirements. Although some services exhibit modest changes, either because demand is already close to being met or because staffing is not the primary constraint, the broader pattern confirms that workforce expansion has system-wide benefits. These additional services represent meaningful opportunities to avert disease burden; for example, earlier detection and treatment through increased HIV-related HSIs could help reduce progression to AIDS^31^.

Increased officer availability in the Target scenario does facilitate the ability to deliver more treatments for conditions directly; however, it still may not translate linearly to a proportional reduction in DALYs, as increased treatment delivery does not necessarily guarantee a reduction in morbidity or mortality. Not all interventions have the same effect on reducing ill health, depending on treatment efficiency, accuracy of diagnostics, and stage of intervention, all of which are captured by the TLO model. In line with the 88.6% shortfall to the Target in Mental officers shown in Fig. 1, addressing this gap would substantially expand officer availability and enable delivery of additional services. For instance, while antidepressants and talking therapies show efficacy in improving depression outcomes (the relative risk of resolution for depression is increased by 1.5 with the use of antidepressants and 1.1 with talking therapies), their ultimate effect on DALYs will rely on the proportion of cases achieving remission and the degree to which reduced symptoms lessen the associated disability burden^32^. With Malaria, many services are delivered by the DCSA cadre^33^, particularly rapid diagnostic tests (RDTs) performed at community or Level 0 facilities (Fig. 7 in ^17^ supports this interpretation). In the Target scenario, DCSA staffing is already at the establishment level (0% shortfall, Fig. 1), reflecting the relative maturity of this cadre under existing programmes. Therefore, expanding the workforce does not translate uniformly across diseases or service areas, and these complex, non-linear relationships emphasise the value of a model such as TLO.

The main assumption taken to undertake this analysis is a rigid healthcare system approach (also explained in ^17,26^) that applies “hard constraints” to HCW utilisation, whereby no level of overtime work and/or task-shifting is allowed, and HSIs must take the time that is specified (based on HCWs’ elicited expectation^17^). It is likely that, in reality, some level of elasticity regarding assumptions takes place, such as informal task-shifting, overtime, or varying patient throughput^27^ may influence actual service delivery, although these processes are difficult to quantify. Similarly, both scenarios do not incorporate unexplained staff absences, meaning daily capabilities may be overestimated to a similar extent. The opposing effects of working overtime and some staff absence create some hesitancy around the DALYs incurred and should be noted. However, because both scenarios rest on the same assumptions regarding overtime, absence, and other unobserved processes, the relative difference between scenarios is less likely to be affected. Given that both scenarios considered make identical assumptions about these constraints, our results should therefore be interpreted as reflecting the health-cost of maintaining 2019 staffing levels versus filling all established positions, rather than an absolute forecast of future DALYs.

Another limitation is that the analysis, although dynamically capturing population growth, assumes no growth in the healthcare workforce numbers over the entire simulation period of (2023-2040), and that consumable availability levels remain constant. These simplifying assumptions were taken for tractability. As the same assumptions are applied in both scenarios, we do not expect them to strongly bias our results.

Another factor not captured by the model is the possibility that understaffing perpetuates a self-reinforcing cycle: too few HCWs increase the health burden, which in turn overloads the system, driving burnout among staff, higher rates of absence, and lower care quality^34^. This reduces service delivery further, increasing the health burden again, and can ultimately drive HCWs to leave, worsening shortages. One 2018 cross-sectional study in Malawi of 520 HCWs involved in HIV care (a cause with potential for substantial DALYs reduction in our analysis (Fig. 3)) 62% met the criteria for burnout, defined as scoring in the mid-to-high range on either the Emotional Exhaustion or Depersonalization subscales of the Maslach Burnout Inventory and burnout was also significantly associated with self-reported suboptimal patient care^35^. Additionally, absenteeism among HCWs in countries across Sub-Saharan Africa has shown rates as high as 50% due to a range of reasons^36,37^, including emotional demand and their health, with far-reaching implications such as causing patients to forego treatment-seeking due to delays ^38,39^. As these factors are not explicitly included in our modelling framework, the relative health gains that could be achieved from fully filling establishment positions could be underestimated.

Finally, our analysis does not address whether filling established posts is the right strategy for health system strengthening. Previous analyses^17,20,40^ suggest that established positions may not correspond well to population health needs. Therefore, the conclusion of this study is not necessarily that government and donors should immediately fill all unfilled posts, but rather that there is a measurable health cost to continued shortfalls under current establishment assumptions. Malawi faces many competing demands on its limited health-sector resources, and we do not compare the health gains from staffing increases with alternative uses of funds. Future analyses should consider HRH expansion paths that are explicitly aligned with the health needs of the population, rather than relying on workforce establishment sizes that may have been based on limited evidence.

## Conclusion

We have found clear evidence of and quantified the lost potential health gains of unfilled health workforce positions in Malawi. The TLO model effectively links healthcare needs with the epidemiological and behavioural patterns within the population, offering a powerful mechanistic means to estimate the DALYs that could be averted through changes in health workforce availability and the establishment of HRH planning. Our findings should provide useful guidance for policymakers in Malawi as they work towards the objectives set out in the HSSP-III and achieve UHC.

## Declarations

### Ethics Statement

The Thanzi La Onse project received ethical approval from the College of Medicine Malawi Research Ethics Committee (COMREC, P.10/19/2820) in Malawi. Only publicly available anonymised secondary data is used in the Thanzi La Onse model; therefore, individual informed consent was not required.

### Consent for publication

Not applicable.

### Availability of data and materials

The Thanzi La Onse model is open source and available for review and usage at https://github.com/UCL/TLOmodel.

### Competing interests

The author’s declare no competing interests.

### Funding

The project is funded by The Wellcome Trust (223120/Z/21/Z to TBH) and contributed to the salaries of BS, REM, SB, TDM and MM; BS, REM, SB, TDM, TBH and MM acknowledge funding from the MRC Centre for Global Infectious Disease Analysis (reference MR/X020258/1), funded by the UK Medical Research Council (MRC). This UK-funded award is carried out in the frame of the Global Health EDCTP3 Joint Undertaking. The funders had no role in study design; in data collection, interpretation, and analysis; in the writing of the report; and in the decision to submit the article for publication. The authors confirmed the independence from funders and had full access to all of the data (including statistical reports and tables) in the study and can take responsibility for the integrity of the data and the accuracy of the data analysis.

### Authors’ contributions

MM, TBH, BS, and AP contributed to the conceptualisation of the study. AP and MM wrote the main manuscript text and AP prepared all the analysis and figures. AP wrote the original draft, MM contributed to the data curation and guidance on visualisation. TBH, BS, and MM accessed and verified the data. All authors reviewed and edited the manuscript.

## Notes

### Competing Interest Statement

The authors have declared no competing interest.

### Funding Statement

Yes

### Author Declarations

Kamuzu University of Health Sciences Research and Ethics Committee (COMREC), has reviewed and approved a study entitled Thanzi La Mawa on 15/11/2023.

